# Cost-effectiveness of the next generation of RSV intervention strategies

**DOI:** 10.1101/19009977

**Authors:** David Hodgson, Richard Pebody, Jasmina Panovska-Griffiths, Marc Baguelin, Katherine E. Atkins

**Author notes:** contributed equally (Jasmina Panovska-Griffiths, Marc Baguelin, Katherine E. Atkins).

## Abstract

**Background:** With a suite of promising new RSV prophylactics on the horizon, including long-acting monoclonal antibodies and new vaccines, it is likely that one or more of these will replace the current monoclonal Palivizumab programme. However, choosing the optimal intervention programme will require balancing the costs of the programmes with the health benefits accrued.

**Methods:** To compare the next generation of RSV prophylactics, we integrated a novel transmission model with an economic analysis. We estimated key epidemiological parameters by calibrating the model to seven years of historical epidemiological data using a Bayesian approach. We determined the cost-effective and affordable maximum purchase price for a comprehensive suite of intervention programmes.

**Findings:** Our transmission model suggests that maternal protection of infants is seasonal, with 2-14% of infants born with protection against RSV. Our economic analysis found that to cost-effectively and affordably replace the current monoclonal antibody Palivizumab programme with long-acting monoclonal antibodies, the purchase price per dose would have to be less than around £4,350 but dropping to £200 for vaccinated heightened risk infants or £90 for all infants. A seasonal maternal vaccine would have to be priced less than £85 to be cost-effective and affordable. While vaccinating pre-school and school-age children is likely not cost-effective relative to elderly vaccination programmes, vaccinating the elderly is not likely to be affordable. Conversely, vaccinating infants at 2 months seasonally would be cost-effective and affordable if priced less than £80.

**Interpretations:** In a setting with seasonal RSV epidemiology, maternal protection conferred to newborns is also seasonal, an assumption not previously incorporated in transmission models of RSV. For a country with seasonal RSV dynamics like England, seasonal programmes rather than year-round intervention programmes are always optimal.

**Funding:** Medical Research Council and National Institute for Health Research

**RESEARCH IN CONTEXT:** *Evidence before this study:* A recent systematic review identified RSV prophylactic candidates currently in clinical trials, including a maternal vaccine (RSV F-nanoparticles vaccine), long-acting monoclonal antibodies aimed at neonates (MEDI8897), and an active adenovirus vector-based vaccine aimed at infants and/or the elderly (ChAd155-RSV). Given the significant observed health burden due to RSV in children, these products, which are mainly aimed at children, are likely to be effective at preventing RSV disease. However, uncertainties surrounding i) the dynamics of maternally-derived protection in neonates, ii) the impact of herd immunity, and iii) the purchasing cost of these prophylactics, means it is not clear if these products are cost-effective. Therefore, evaluating the purchasing price required for these prophylactics to remain cost-effective using a dynamic transmission model, in which the herd immunity is included and the dynamics of maternally-derived immunity is determined by calibrating the model to data, is a public health priority.

*Added-value of this study:* Our study finds that in a setting with seasonal RSV epidemiology, maternal protection conferred to newborns is also seasonal. In addition, our study estimates the maximum purchasing price per course for various potential RSV intervention programmes to be cost-effective in England, assuming a cost-effectiveness threshold of £20,000/QALY as recommended by the National Institute of Clinical Excellence (NICE). We find that to cost-effectively and affordably replace the current monoclonal antibody Palivizumab programme with long-acting monoclonal antibodies, the purchase price per dose would have to be less than around £4,350 but dropping to £200 for vaccinated heightened risk infants or £90 for all infants. A seasonal maternal vaccine would have to be priced less than £85 to be cost-effective and affordable. While vaccinating pre-school and school-age children is likely not cost-effective relative to elderly vaccination programmes, and vaccinating the elderly is not likely to be affordable.

*Implications of all the available evidence:* The seasonal protection conferred to newborns is consistent with empirical immunological data from maternal cord blood and ecological evidence from hospital records. Further, extending a monoclonal antibody programme would be possible if there is a considerable drop in price and maternal vaccination remains the only realistic vaccination strategy in the UK. With further clinical trials planned for RSV F-nanoparticles vaccine to evaluate its efficacy, and with clinical trial completion dates set at the end of 2021 for the other two prophylactic candidates, the results of this study provide a comprehensive overview of the impact of potential RSV intervention programmes both in the present and in the coming years.

## Introduction

Respiratory Syncytial Virus (RSV) is the most common cause of acute lower respiratory infection in children under five years of age globally, causing 48,000-74,500 deaths annually.^1^ The sole pharmaceutical prevention strategy, a monoclonal antibody (Palivizumab), is costly and only available to infants in high-income countries and only to those at most risk of RSV-related complications.^2^ This gap in prevention strategies leaves the majority of infants vulnerable to infection.

There are currently over 40 RSV prophylactic candidates in pre-clinical or clinical trials,^3^ those furthest along in development include long-acting monoclonal antibodies (e.g. MEDI8897 by *MedImmune*),^4^ and maternal, childhood, and elderly vaccines (e.g. RSV F-nanoparticle vaccine by *Novavax*, ChAd155-RSV, by *GlaxoSmithKline* and Ad26.RSV.preF by *Jensen* respectively).^5,6^ Although missing its primary endpoint, a recent Stage III trial of the *Novavax* RSV F-nanoparticle vaccine showed promising results, preventing RSV-related lower respiratory tract infections and hospitalisations in babies born to vaccinated mothers in the South Africa site.^5^ While, Stage II trial results suggest that the *MedImmune* MEDI8897 long-acting monoclonal antibodies are effective at preventing RSV-disease in neonates for at least 150 days post-administration—five times longer than a single dose of Palivizumab.^4^ Stage II trial results for the adenovirus vectored vaccines *GlaxoSmithKline* ChAd155-RSV and *Jensen* Ad26.RSV.preF suggest that they are well tolerated and safe in their respective target groups of infants and the elderly respectively, though we currently lack efficacy results.^6^

Deciding which, if any, of this suite of pharmaceutical prophylactics to adopt requires an integrated approach in which all the health benefits accrued by targeted specific subpopulations (intervention strategies)—both by direct and indirect protection and across all ages—can be accurately compared. Moreover, with multiple new prophylactics likely to arrive to license at a similar time, understanding the relative efficiency of potential intervention strategies at controlling RSV burden, and therefore what we should be willing to pay for them, will dominate decision-making on future RSV intervention strategies.

In this study we developed such an integrated approach by combining a novel age-stratified epidemiological transmission model for RSV into a cost-effectiveness framework. The model was calibrated using a Bayesian inference framework to seven years of RSV incidence data from England. The cost-effectiveness analysis was undertaken according to the National Institute of Clinical Excellence (NICE) reference case.^7^ Using this approach, we were able to determine the maximum purchasing prices for the next generation of RSV intervention strategies to be cost-effective and affordable.

## METHODS

### RSV model structure

We modelled the number of individuals in six different epidemiological states (*M, S, E, I, A* and *R*). When a susceptible individual (*S*) acquires infection, they move to an exposed but not infectious state (*E*) for an average of 1/σ days, after which they become infectious with either symptomatic (*I*) or asymptomatic (*A*) infection. After an infectious period of 1/γ days, individuals move to a protected state (*R)* for a period of 1/ω days, after which they become susceptible to reinfection (*S*). We assume that only babies born to mothers who have recently been infected with RSV and who therefore have high levels of antibody (and thus, in state *R*) are maternally protected (*M*) for a period of 1/ξ after birth, with the remaining babies born susceptible to infection (*S*) (**Supplementary Information 1 Figure S1**). We tested this assumption to the alternative where all babies are born with temporary maternal immunity, similar to previous models (e.g. Kinyanjui et al.^8^) using the deviance information criterion (DIC) (**Supplementary Information 1 Section 1.1, Figure S2**). We stratified individuals into 25 age groups (monthly up to 11 months of age, and then 1, 2, 3, 4, 5–9, 10–14, 15–24, 25–34, 35–44, 45–54, 55–64, 65– 74, 75+ years) and also tracked the number of individuals had experienced zero, one, and two or more previous infections (denoted by the subscripts 0, 1, 2, 3). Consistent with empirical data, we assume that the proportion of individuals who experienced asymptomatic infection is dependent on age^9^ and the duration of infection and susceptibility to infection are dependent on the number of previous RSV infections.^10,11^ We assume that the contact rate between two age groups is proportional to the mean number of daily physical and conversational contacts made between those age groups—parameterised using empirical data from England and Wales (**Supplementary Information 1 Section 1.2**).^12,13^ We captured the strongly seasonal dynamics of RSV in temperate climates by multiplying the per-contact transmission rate with a seasonal forcing term (**Supplementary Information 1 Section 1.3)**.

To capture the current impact of administering Palivizumab, we tracked very high risk (VHR) infants aged between 0-8 months who are eligible for a course of Palivizumab during the RSV season, that is those born premature with Chronic Lung Disease or Congenital Heart Disease during the months of October to February, the RSV season).^2^ For these VHR infants, we assumed that 90% receive Palivizumab with 33·8% acquiring immediate protection which lasts for an average of 1/ω_pal_ = 150 days, after which they return to the primary susceptible compartment (*S*_0_).^14^

### Model parameterisation and calibration

We used a Bayesian Markov chain Monte Carlo (MCMC) approach^15^ to fit the model to the confirmed number of positive weekly RSV samples in England collected via the Respiratory DataMart System (RDMS) between July 2010 and June 2017.^16^ We used a binomial likelihood function that assumes an age-specific reporting rate of RSV positive samples. To estimate how the reporting rates of RSV infection varied across ages, we tested five different assumptions about the age stratification (number and age grouping) using the Deviance Information Criterion (DIC) (**Supplementary Information 1 Section 3, Figure S5, Table S4**). We constructed the prior distributions for all epidemiological parameters after a comprehensive synthesis of the literature (**Supplementary Information 1 Section 2, Figures S4 and Tables S3**).^9–11,17–26^ The output of the calibration is a joint posterior distribution for all the fitted parameters of the transmission model. To compare our model to the weekly number of RSV positive samples we multiplied the model-predicted weekly incidence of symptomatic cases with the fitted age-specific reporting rates.

### Intervention model structure

#### Status quo

We assume that Palivizumab is currently administered to 90% of VHR infants at birth between October to February inclusive (PAL-VHR-S, **Supplementary Information 1 Section 4 Figure S6**). We compare this status quo to the following three alternative intervention strategies.

#### Long-acting monoclonal antibodies

We tracked the number of infants protected by long-acting monoclonal antibodies, *V*_M_, who remain protected after birth for an average of 1/ω_mab_ = 250 days after which they return to *S*_0_ (**Supplementary Information 1 Section 4 Figure S7**). However, we relaxed this assumption in an uncertainty analysis. We evaluated three seasonal programmes that administer a single dose of long-acting monoclonal antibodies at birth i) to those who are currently eligible for Palivizumab (MAB-VHR-S), ii) to both VHR infants and infants who are at heightened risk (HR) of developing complications due to respiratory disease (MAB-HR-S), iii) to all infants regardless of risk (MAB-ALL-S). We evaluated two additional seasonal programmes that extend administration (iv) to all VHR and high-risk (HR) infants under six months (MAB-HR-S+) and v) to all infants under six months (MAB-ALL-S+) throughout October only.^6^ We assume that these programmes would replace the existing Palivizumab programme; that they all achieve the same coverage as Palivizumab and that the efficacy per course is 70·1% (95% Confidence Interval (CI) 52·3–81·0%).^4,27^

#### Childhood/elderly vaccination

We assumed that a single dose of a vaccine conferred the same protection as that of a natural infection, such that 83·0% (95% CI 75·0–88·0%) of vaccinated individuals in the *i*th previous infection group who are susceptible (*S*_i_) are moved to respective recovered group (*R*_i_) after a delay reflect the build up of antibody immunity (**Supplementary Information 1 Section 4 Figure S8**).^28^ We considered two vaccination programmes aimed at infants aged 2 months old; one administered seasonally (VAC-INF-S) and one year-round (VAC-INF-A), both achieving a coverage of 90%, consistent with the DTaP/IPV/Hib/HepB/PCV/Rota primary series vaccination coverage in England. We also considered two seasonal vaccination programmes aimed at elderly persons: one for those aged 75 years and older (VAC-75-S) and one for those aged 65 years and older (VAC-65-S), both achieving a coverage of 70%, consistent with vaccination coverage for the elderly influenza vaccine programme.^29,30^ Finally, we considered three seasonal programmes aimed at preschool children (aged 2–4 years, VAC-2-4-S) and school-age children (aged 5--9 years, VAC-5-9-S, and aged 5–14 years, VAC-5-14-S), that achieve a coverage of 45% and 60% respectively, consistent with the live attenuated influenza vaccination programme in England.^29^ We assumed that all these vaccine programmes would be administered in addition to the existing Palivizumab programme in the UK.

#### Maternal vaccination

To evaluate the direct effect on infants of vaccinating pregnant women, we used the results of Novavax’s maternal vaccine Stage III trial that found 41·4% (95% CI 4·1– 64·2) of infants born to these mothers are protected against infection for the first 3 months of life (**Table 2**).^5^ Consistent with the trial, we assume pregnant women are vaccinated at any point between 28 and 32 weeks gestation (**Supplementary Information 1 Section 4 Figure S9**).

**Table 2.** Intervention model parameters. CrI—credible interval. Fitted distributions

|  | Parameter | Mean value (95% CI where applicable) | Reference |
| --- | --- | --- | --- |
| *Palivizumab* |  |  |  |
|  | Delay between administration and protection (days) | Immediate (Fixed) | 14 |
| $\omega_{\text{pal}}$ | Average period of protection (days) | 150 (fixed) | 14 |
| $e_{\text{pal}}$ | Efficacy on VHR infants (%) | 33.8 (0.0–66.6)<sup>1</sup> | 14 |
| *Long-acting monoclonal antibodies* |  |  |  |
|  | Delay between administration and protection (days) | Immediate (Fixed) | 4 |
| $\omega_{\text{mab}}$ | Average period of protection (days) | 275 (fixed) | 4 |
| $e_{\text{mab}}^{\text{S}}$ | Efficacy against symptomatic infection (%) | 70.1 (52.3–81.0)<sup>2</sup> | 27 |
| $e_{\text{mab}}^{\text{H}}$ | Efficacy against hospitalisation (%) | 78.4 (51.9–90.3)<sup>3</sup> | 27 |
| *Childhood/elderly vaccine* |  |  |  |
| $d_{\text{vac}}$ | Delay between administration and protection (days) | 11.4 (2.8–22.1)<sup>4</sup> | 28 |
| $\omega$ | Average period of protection (days) | Same as post-infection immunity ($1/\omega$) | 28 |
| $e_{\text{vac}}$ | Efficacy against all infections (%) | 83.0 (75.0–88.0)<sup>5</sup> | 28 |
| *Novavax vaccine* |  |  |  |
| $d_{\text{mat}}^2$ | Average period of protection (days) | 133.5 (119.6–146.1) | Same as maternally derived immunity |
| $e_{\text{mat}}^{\text{S}}$ | Efficacy against symptomatic infection (%) | 41.4 (4.1–64.2)<sup>6</sup> | 5 |
| $e_{\text{mat}}^{\text{H}}$ | Efficacy against hospitalisations (%) | 53.5 (23.0–71.9)<sup>7</sup> | 5 |

To evaluate the indirect effects of maternal vaccination while maintaining computational tractability and epidemiological realism, we used a previously published method for evaluating the impact of parental vaccination.^31^ In brief, this method tracks the number of mothers of infants less than one year of age, and the number of these women who are participating in a maternal vaccination programme. The contact rate between mothers and their children is explicitly modelled using the number of household and non-household contacts, as reported by the Great Britain arm of the POLYMOD study.^12,13^ Accordingly, the force of infection between mothers and their infants is updated to reflect the vaccination status of the mother. We assume that the vaccinated mothers are themselves temporarily protected from infection consistent with the protection afforded by the childhood/elderly vaccination assumptions above (**Supplementary Information 1 Section 4 Figure S9**). We considered two maternal vaccination programmes, which are given in combination with the existing Palivizumab programme: a seasonal programme (MAT-S), and one administered year-round, (MAT-A) with a coverage of 60% as observed for prepartum Tdap vaccination in England.^5,29^

#### Optimising seasonal administration

To allow an unbiased comparison of the seasonal programmes, our framework assumes the programmes are given continuously for five months. For programmes that administer Palivizumab and long-acting monoclonal antibodies, we assume administration occurs during the Palivizumab-recommended time period of October to February. To determine the period of administration for the remaining intervention programmes, we chose the five-month period that resulted in the largest QALY gain relative to status quo.

### Economic model

#### Clinical outcomes

For each intervention strategy, the economic model estimated the number of cases averted for five different RSV-associated clinical outcomes: symptomatic infection, GP consultations, hospital admissions, hospital bed days and deaths. The number of symptomatic cases averted is estimated directly from the transmission model. To estimate the number of cases averted for the remaining four outcomes, we first calculated the per-infection probability that an individual experiences each clinical outcome by dividing the reported annual incidence rates for each outcome taken from previous burden studies in England^32–38^ (**Supplementary Information 1 Section 5.1, Figure S10)** by the transmission-model-estimated annual incidence for RSV under the status quo. Then, to calculate the number of cases averted for each outcome under each intervention strategy, we multiplied the estimated number of RSV-cases averted from the intervention model by the per-infection probability of each outcome.

#### Quality of life loss

In line with our previously estimated quality-adjusted life year (QALY) loss estimates per RSV episode for England, we assume that each GP consultation or hospitalisation resulted in a QALY loss of 4·098 × 10^−3^ (0·624 × 10^−3^–13·141 × 10^−3^) and 2·990 × 10^−3^ (0·346 × 10^−3^–11·387 × 10^−3^) for under fives and over fives respectively, while other symptomatic non-healthcare seeking infections resulted in a QALY loss of 2·336 × 10^−3^ (95% CI 0·269 × 10^−3^–9·255 × 10^−3^) and 1·448 × 10^−3^ (95% CI 0·135 × 10^−3^–5·928 × 10^−3^).^39^ QALY loss due to death was commensurate with the remaining number of expected healthy years of life remaining in the individual (**Supplementary Information 1 Section 5.2)**.

#### Costs

Costs were calculated in 2018 GBP, from the perspective of the NHS. The cost per GP consultation was calculated by assuming an average GP consultation time of 9 minutes at a cost of £4.00 a minute (£36.00).^40,41^ The cost per hospital bed day for children less than five years of age was calculated using the non-elective costs for paediatric Bronchitis (Health Resource group (HRG) PD15A–D)—the main cause of RSV-associated hospitalisations.^42,43^ The cost per hospital bed day for children five years and older was determined using the non-elective costs for unspecified Acute Lower Respiratory Infection (HRG DZ22K–Q).^43^ We assumed maternal, infant and elderly vaccines take 15 minutes to administer in a GP clinic at a cost of £9 per course (assuming one dose per course).^41^ Similarly, we assumed long-acting monoclonal antibodies and Palivizumab, take 15 minutes to administer in hospital by a nurse at a cost of £11.50 per course for long-acting monoclonal antibodies and £57.50 per course (5 doses) for Palivizumab.^41^ A course of Palivizumab costs £4035 (5 doses at £807 each).^43^

#### Cost-effectiveness analysis

We conducted three separate cost-effectiveness analyses. First, we calculated the incremental cost-effectiveness ratio (ICER) of replacing the Palivizumab with any of the long-acting monoclonal programmes (MAB-VHR-S, MAB-HR-S, MAB-HR-S+, MAB-ALL-S, and MAB-ALL-S+). Second, we calculated the ICERs of supplementing the Palvizumab programme with the childhood or elderly vaccine programmes (VAC-INF-S, VAC-INF-A, VAC-2-4-S, VAC-5-9-S, VAC-5-14-S, VAC-75-S, VAC-65-S). Third, we calculated the ICER of supplementing the Palivizumab programme with the maternal vaccine programmes (MAT-S, MAT-A). For each of these three cost-effective analyses, using the non-dominated programmes only, we calculated the maximum price per course that would make each strategy cost-effective, assuming a cost-effectiveness threshold of £20,000/QALY (**Supplementary Information 1 Section 5.3**). All costs and effects were discounted at a rate of 3.5% over a 10-year time horizon.^7^ For each intervention strategy, we calculated the credibility intervals using 1,000 Monte Carlo samples. For each Monte Carlo sample, we first estimated the number of RSV cases averted over the time horizon per outcome for an intervention strategy by sampling from the joint posterior distribution and running the intervention model for 10 years. Then, by sampling from the per-infection probability of each outcome occurring, we converted the number of RSV cases averted to the number of outcomes averted. Finally, we combined sampled values from the cost distributions with the number of each clinical outcome averted to calculate the distribution of the maximum price per prophylactic course.

#### Affordability

An intervention strategy is considered affordable if it costs less than £20 million annually during the first three years of implementation.^44^ Using this definition, we calculated the affordable purchasing price per course for each non-dominated programme, by subtracting the total, undiscounted cost of administering the intervention strategy for the first three years from £60 million (3 years at £20 million each) and dividing by the total number of courses given during this period.^44^

#### Calculations and code

The model was programmed in C++ with the code available at https://github.com/dchodge/rsv_trans_model. The figures were generated in Mathematica version 11.0.0.^45^

## RESULTS

### RSV Epidemiology

Our model comparison analyses suggested that maternal immunity was conferred seasonality according to the prevalence of recently infected pregnant mothers (**Supplementary Information 2 Figure S1**). Furthermore, we found that there is a likely exponential decrease in the reporting rates between the ages of 0-5 years, and fixed reporting rates for 5-55 years and 55 years and over (**Supplementary Information 2 Figure S2**).

The model reproduces the age distribution of RSV incidence (**Figure 1a-c, Supplementary Information 2 Figure 3–4**). Using the calibration method, we are able to estimate parameters that have been difficult to evaluate directly from epidemiological studies (**Supplementary Information 2 Figure 5)**. First, our model predicts that between 68-81% of infants experience an RSV infection in their first year of life, with subsequent infection risk generally decreasing with age (**Figure 1d**). Deviations away from this decreasing trend occur in age groups which have the highest number of daily contacts (**Figure 1d**). Second, we estimated the average duration of maternal immunity and post-infection immunity as 134 days (95% CrI 120–146) and 359 days (95% CI 351–365), respectively. Third, the model estimated that asymptomatic infections are 63% (95% CrI 54%–72%) as infectious as symptomatic infections **(Table 1)**, and finally, we found that 14% of babies are born with protection to RSV during March, compared with 2% prior to the RSV season in September and October (**Supplementary Information 2 Figure 2)**.

**Table 1.** Posterior distributions of the model parameters used in the transmission model of RSV. CrI— credible interval.

|  | Parameter | Mean value ( 95% CrI of posterior where applicable) | Reference for fixed value or prior distribution |
| --- | --- | --- | --- |
| $\mu$ | Daily number of live births | 1863 (Fixed) | 20 |
| $1/\xi$ | Average duration of maternally-derived immunity (days) | 133.5 (119.6–146.1) | 19,22,24 |
| $1/\omega$ | Average duration of post-infection immunity (days) | 358.9 (350.7–364.7) | 25,26 |
| $1/\sigma$ | Average duration of exposure (days) | 4.98 (4.54–5.37) | 18 |
| $1/\gamma_0$ | Average duration of primary infection (days) | 6.16 (5.68–6.63) | 11 |
| $g_1$ | Decrease in secondary infection duration relative to primary | 0.87 (0.83–0.91) | 11 |
| $g_2$ | Decrease in subsequent infection duration relative to secondary | 0.79 (0.73–0.86) | 11,18 |
| $p^{<1}$ | Proportion asymptomatic (<1 years) | 0.0916 (0.031–0.158) | 9 |
| $p^{1-4}$ | Proportion asymptomatic (1–4 years) | 0.163 (0.092–0.223) | 9 |
| $p^{5-14}$ | Proportion asymptomatic (5–14 years) | 0.516 (0.460–0.572) | 9 |
| $p^{15+}$ | Proportion asymptomatic (15+ years) | 0.753 (0.656–0.829) | 9 |
| $\alpha$ | Relative reduction in infectiousness for asymptomatic infections | 0.634 (0.541–0.724) | Fitted |
| $q_p$ | Probability of RSV transmission per physical contact | 0.0972 (0.093– 0.099) | Fitted |
| $q_c$ | Relative reduction in probability of RSV transmission per conversational contact compared to physical contact | 0.998 (0.996– 1.000) | Fitted |
| $b_1$ | Relative amplitude of transmission during peak | 1.998 (1.992–2.000) | Fitted |
| $\varphi$ | Seasonal shift in transmission | 0.614 (0.607–0.624) | Fitted |
| $\psi$ | Seasonality wavelength constant | 0.236 (0.220– 0.252) | Fitted |
| *Susceptibility* |  |  |  |
| $\delta_1$ | Secondary infection (relative to primary) | 0.89 (0.85-0.93) | 10 |
| $\delta_2$ | Tertiary infection (relative to secondary) | 0.81 (0.74-0.85) | 10 |
| $\delta_3$ | Subsequent infections (relative to tertiary) | 0.33 (0.31-0.37) | 10 |
| *Probability that an RSV infection is reported* |  |  |  |
| $\epsilon^j$ | 0-4 years | $\exp(-4.602 - 0.233j)$ | 10,23 |
| $\epsilon^{17}$ | 5--54 years | 0.0000305, (0.0000290-0.0000320) | 0,23 |
| $\epsilon^{18}$ | 55+ years | 0.000174,<br>(0.000134-0.000160) | 0,23 |

**Figure 1.**
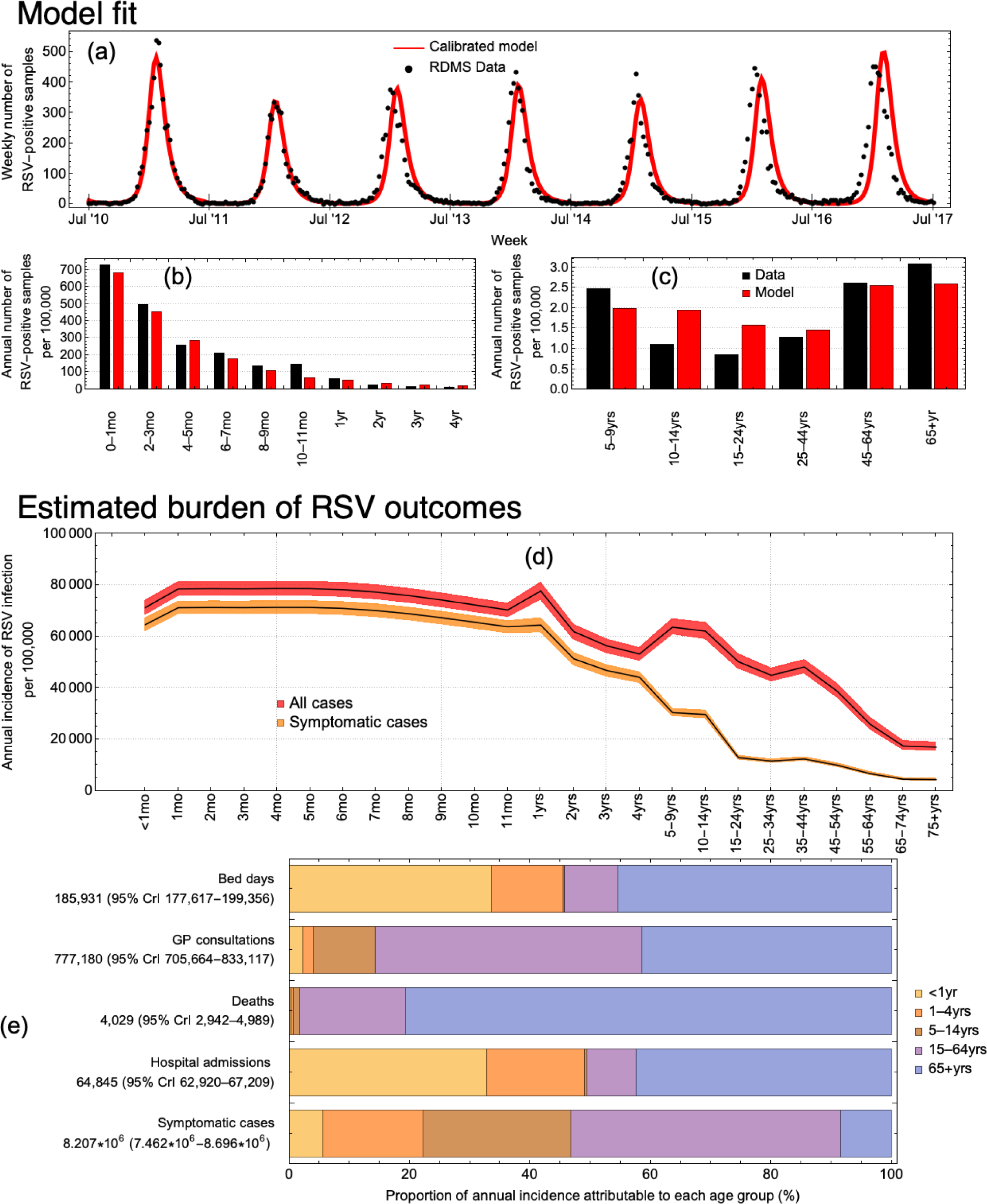
The calibrated model and the incidence of RSV-associated outcomes. (a) The model-estimated mean number of new weekly infections fit to the reported RSV-positive samples from July 2010 until July 2017 in England. (b,c) Model-predicted mean annual number of new infections per age group with the reported RSV-positive samples. (d) The model-predicted incidence of any and symptomatic RSV infections. (e) Age group attribution to each health care outcome.

**Figure 2.**
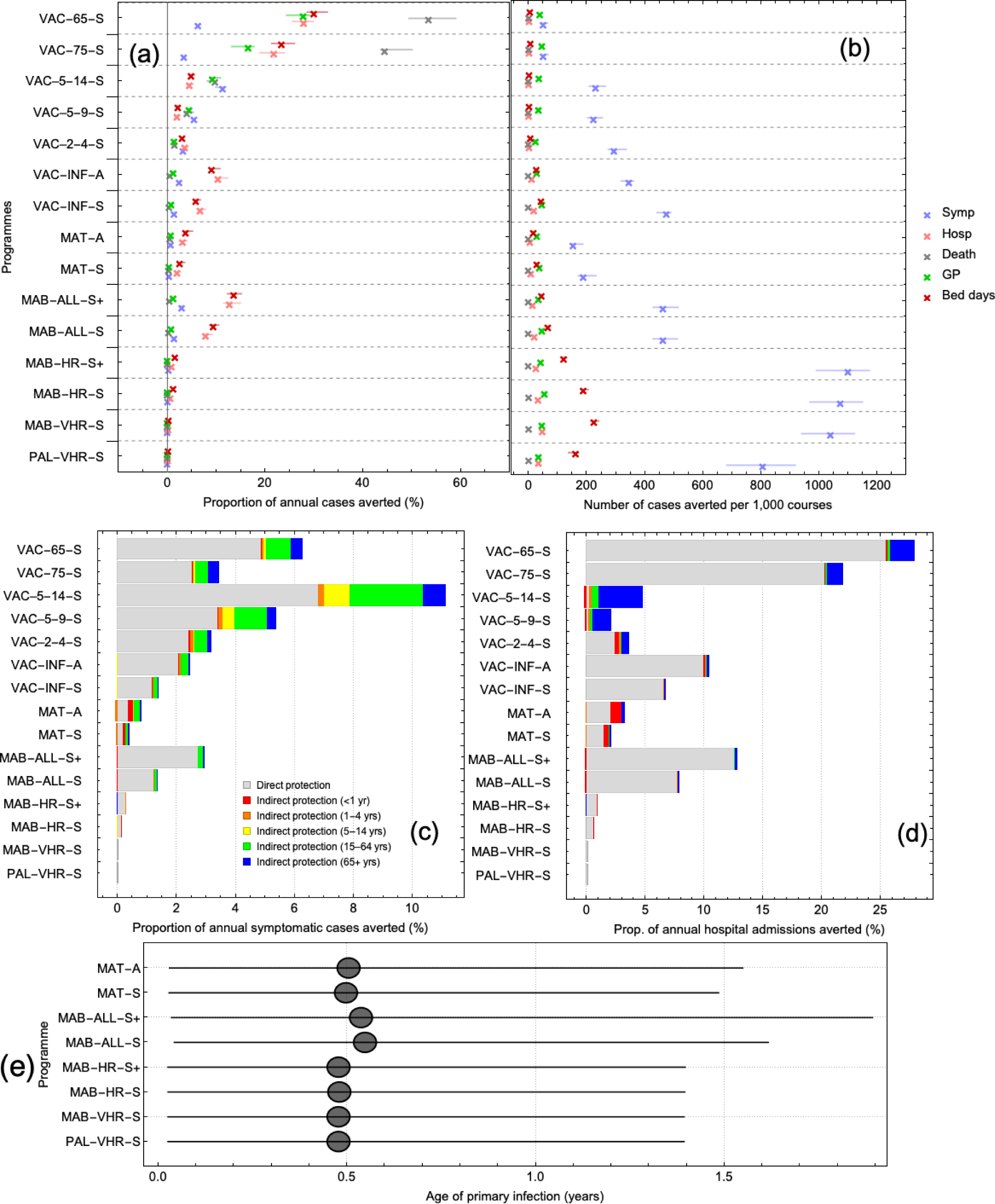
The impact of the 14 intervention programmes. (a) Total effectiveness (direct and indirect effects) of each intervention programme at preventing five healthcare outcomes (symptomatic infection, hospital admission, death, GP consultations, and bed days). (b) Efficiency of programmes. (c-d) Effectiveness of each intervention strategies in terms of direct (gray) and indirect effects for symptomatic infection (c) and hospitalised cases (d). (e) Median age of primary infection for long-acting monoclonal antibodies and maternal vaccines.

**Figure 3.**
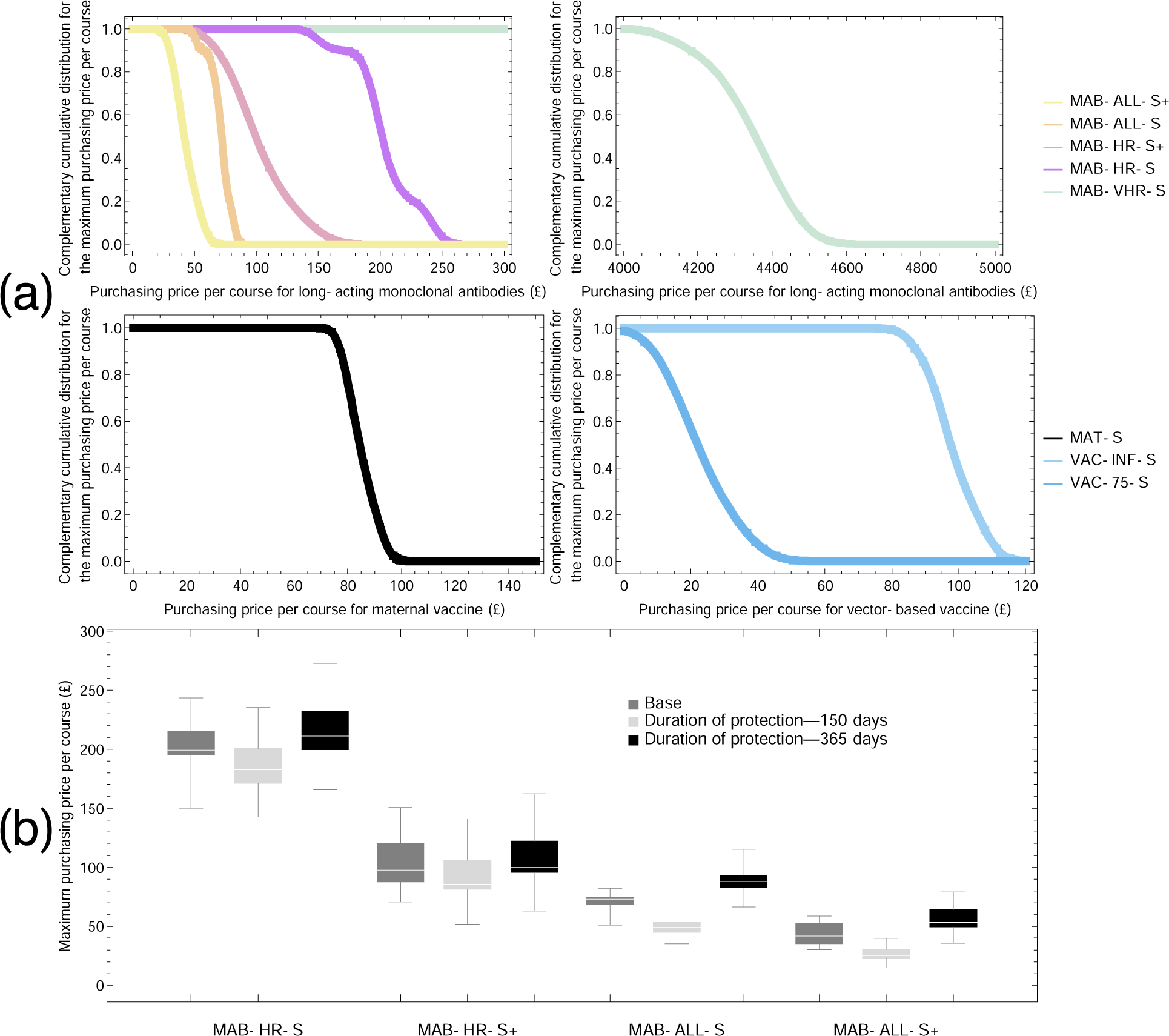
Maximum purchase price for a course of treatment to remain cost-effective assuming a cost-effectiveness threshold of £20,000 QALY. (a) Probability of cost-effectiveness for each of the non-dominated programmes over a range of purchasing prices. (b) Sensitivity analysis on the duration of protection for the monoclonal antibodies and its effect on the maximum purchasing price per course.

### Probability of clinical outcomes

The average probability of consulting a GP due to RSV infection is highest in children less than 5 years of age (0·006–0·065) and adults 65 years and older (0·103–0·132). The average probability of death per-infection is highest in adults over 75 years (0·002) and rare in children and other adults in the remaining age groups (less than 3 in every 100,000 infections). The average probability of hospitalisation is highest in infants below 1 year of age (0·010–0·097), with peak risk occurring at 1 month of age, and lowest risk in persons aged 5-45 years of age (less than one in every 10,000 infections). HR and VHR infants have an increased risk of hospitalisation of 0·0138–0·129 and 0·14–0·37 respectively, compared with other infants (0·010–0·097). Similarly, the average number of bed days experienced per hospitalisation is greatest in infants less than 1 year of age (1–5) with the longest stays occurring at 1 month of age, and HR and VHR infants seeing an increase in the number of bed days of 5–7 and 8–25 respectively. (**Supplementary Information 2 Figure 6**).

### Impact of intervention strategies

#### Long-acting monoclonal antibodies

The seasonal programmes aimed at VHR infants or VHR and HR infants (MAB-VHR-S and MAB-HR-S, respectively) are the most efficient at preventing RSV hospitalisations, preventing 51 (95%CI 43–55) and 36 (95% CI 30–39) hospital cases per 1,000 administered courses. These intervention programmes are not effective in raising the median age of primary infection. (**Figure 2b**).

#### Childhood/elderly vaccination

We found that to maximise the health benefit of the seasonal vaccination programmes, the optimal period of administration is between November and March for elderly programmes, October to February for the VAC-2-4-S and VAC-5-9-S programmes, and August to December for the VAC-5-14-S programme. (**Supplementary Information 2 Figure 7**). Vaccinating individuals 65 years and over is the most effective programme at preventing the total number of GP consultations, hospitals, bed days and deaths (23%, 25%, 26% and 49% reductions respectively) (**Figure 2a**). However, the large size of the target group means this programme is inefficient, preventing 19·03, 1·63, 4·34, and 0·25 cases of GP consultations, hospitals, bed days and deaths respectively per 1,000 vaccine courses. The most effective school-age programme is the 5–14 year old programme, preventing 4·5% (95% CrI 3·9–5·4) of hospitalised cases. School-age programmes confer considerable herd protection, with 91.5% of the 5-9 year old programme and 94·9% of 5-14 year old programme of averted hospitalised cases due to indirect protection (**Figure 2c, Supplementary Information 2 Figure 8**).

#### Maternal vaccination

Our results suggest that, to maximise the health benefit for a seasonal third trimester maternal programme, the optimal period of administration is from August until December (**Supplementary Information 2 Figure 7)**. Such a programme prevents 8·5 (95% CrI 7·4–10·3) hospitalised cases per 1,000 vaccine courses administered, with 22-30% of the hospitalised cases prevented in infants less than 1 year of age attributable to indirect protection from vaccinated mothers (**Figure 2d**). Though the seasonal maternal programme is more efficient than its year-round counterpart, it is less efficient at preventing hospitalised cases than any of the long-acting monoclonal antibodies programmes.

### Cost-effectiveness analysis of intervention strategies

#### Long-acting monoclonal antibodies

The maximum purchasing price per course for the long-acting monoclonal antibodies programme to be cost-effective when administered seasonally to only the VHR infants is £4,342.97 (95% CrI £4,126.31–4,462.25) (**Figure 3**). For this seasonal programme to remain cost-effective after extending to HR neonates (MAB-HR-S), and then to all HR infants less than 6 months at the start of season (MAB-HR-S+), requires substantially lower maximum purchasing prices per course of £201.15 (95% CrI £149.61–243.42) and £87.03 (95% CrI £64.80–116.99) respectively (**Figure 3a**). If the duration of protection varies between 150 and 365 days, the maximum purchasing price for the MAT-HR-S programme would also vary between £185.79-215.02, respectively (**Figure 3b**).

#### Maternal vaccination

The year-round maternal vaccination programme was dominated by the seasonal strategy. The maximum purchasing price per course for the seasonal maternal vaccination to be cost-effective is £85.27 (95% CrI £77.79–93.80) (**Figure 3a**).

#### Childhood/elderly vaccination

The year-round vaccine programme aimed at infants 2 months of age is dominated by its seasonal counterpart, while the 65 years and over programme is dominated by the 75 years and older programme. Further, the pre-school, and school-age programmes are subject to extended dominance by the 75 years and older programme. For the seasonal vaccine programme aimed at infants aged 2 months of age, the maximum purchasing price per course to remain cost-effective is £94.76 (95%CrI 89.09–99.24). Targeting those aged 75 years and older requires a lower purchasing price per course of £20.71 (95% CrI 10.32– 34.64) (**Figure 3a**).

#### Affordability

The long-acting monoclonal antibodies programmes: MAB-VHR-S, MAB-HR-S, and MAB-HR-S+ and the seasonal maternal programme (MAT-S) are affordable if implemented for a cost-effective purchasing price per course (affordable thresholds are £9,395.75, £1,712.46, £873.08, and £121.02 respectively). The seasonal infant programme aimed at 2-month-olds and the 75 years and older programme are affordable if implemented for £79.62 and £3.63 respectively—81% and 16% of the estimated mean maximum purchasing price per course.

## DISCUSSION

This study used a mathematical modelling approach calibrated to seven years of RSV incidence data to evaluate RSV epidemiology and surveillance in a developed country. Integrating this model into a cost-effectiveness framework, we evaluated the likely maximum dose prices of the new generation of RSV preventive pharmaceuticals to make them cost-effective and affordable in England. Our epidemiologic analysis found that maternal protection for infants is likely seasonal, with more babies born with protection against RSV towards the end of the RSV season in March. Our economic analysis found that replacing the existing seasonally administered Palivizumab programme with long-acting monoclonal antibodies would be cost-effective and affordable at a maximum course price of £4403 (95% CrI 4338–4511). Extending the programme to heightened risk or all infants would remain cost-effective and affordable at approximately £200 and £90, respectively. A seasonal maternal vaccination programme would be cost-effective and affordable with a maximum purchasing price per course of £85 (95% CrI 79–91).

This is the first study to use a dynamic transmission model to evaluate how Palivizumab, monoclonal antibodies, and maternal vaccines impact the incidence of RSV-related healthcare outcomes within a single framework. Consequently, this model gives a comprehensive overview of the impact of all currently proposed RSV programmes. This study is also the first to directly link the impact of potential programmes from a dynamic transmission model to a cost-effectiveness analysis (CEA) according to the NICE reference case—the gold standard approach for CEA in England and Wales, and the first to use EQ-5D-based QALY estimates for RSV. The CEA accounts for both the direct and indirect effects of intervention strategies. This approach is of particular importance when comparing the health benefits of vaccinating school-age children through which all reductions in hospitalisations are through indirect protection of other high-risk groups, with those of providing direct protection to newborns where all averted cases are in the newborns themselves.

Our model is the first to test the hypothesis that maternal protection to newborns is seasonal, contrary to the routine assumption in models in which all babies are born with protection to RSV. We found evidence for our alternative hypothesis. This seasonal change in the number of protected new-borns could provide an explanation for previous findings that hospitalisation rates increase for babies born at the start of the RSV season, when our model predicts the lowest fraction of maternally-protected infants, and infants born at the end of the RSV season when the maternal protection is highest experience the lowest risk of RSV-related hospitalisation.^46^ Epidemiological evidence for seasonal changes in maternal protection has also been provided in studies looking at seasonal changes in RSV-specific antibody level from cord titres at birth.^17^ As cord titre influences the rate of severe infections in the first year of life,^24^ seasonal changes could indicate temporal vulnerability in the infant population.

The maternal vaccine in this model is based on Novavax ‘s RSV F-nanoparticle formulation. Recent stage III trial results for this product failed to meet its primary end point of 40% efficacy against RSV lower respiratory tract infections (LRTI) during the first three months of life across all trial sites. However, variations in efficacy were observed depending on region and gestation age at administration. Regional variation in efficacy saw South Africa with promising efficacy estimates of 57% (95% CI 33%–73%) against RSV LRTI, whereas the US site saw no evidence of efficacy.^5^ Although in this analysis, we assume the efficacy of the maternal vaccine is as estimated across all sites, we acknowledge that care should be taken when these results are projected onto the UK, which experiences seasonal RSV similar to the US trial site. Efficacy was also found to vary with gestational age at administration, with vaccination at the start of the third trimester (28-32 weeks) experiencing an efficacy of 54% (95% CI 23%–72%) against RSV-associated hospitalisation and showing superior antibody transfer when compared to administration later in the third trimester (efficacy of 26% (95% CI −23–56). In our study we have chosen the efficacy given at 28-32 weeks gestation as the health care delivery system in England is such that specific uptake periods are feasible in GP clinics if individuals are notified at the relevant time. However, uptake during this specific window may be less feasible in countries with differing healthcare policy and thus lower coverage rates may be observed than used in this study.

Though the results of this analysis suggest that the long-acting monoclonal antibodies and maternal programmes are cost-effective, implementation of these programmes will present clinical and logistical challenges that this analysis has not considered. For example, we assume the same administration price per dose for all the monoclonal antibody programmes. However administration of monoclonal antibodies to the under 6 months, rather than just newborns, will likely be more expensive and achieve lower rates of uptakes, all else equal, as they will need to make a separate appointment at a GP or hospital setting for dose administration. Consequently our results may overestimate the impact and cost-effectiveness of these programmes. Further, in estimating the per-infection risk for RSV-related outcomes, there were no clinical-risk-specific estimates for death and for GP consultations available in the literature, meaning the probability of these outcomes occurring may be underestimated in VHR or HR infants, implying costs and QALY burden of some of the intervention strategies may be conservative. Further studies which help estimate the burden of specific outcomes in England would help reduce uncertainty and increase the accuracy of the model predictions.

In this study, we have used a Bayesian approach to synthesise existing epidemiological and clinical information to estimate the uncertainty in the model parameters and to incorporate uncertainty arising from these parameter estimates to help inform decision-makers about the implementation of new RSV intervention strategies. Our analysis finds that, regardless of the intervention strategy, seasonal administration of a programme is always optimal. Moreover, we found little evidence that strategies aimed at children 2 years and older and those targeted at the elderly would be cost-effective or affordable, respectively. In contrast, long-acting monoclonal antibodies and maternal vaccines may be a cost-effective replacement or addition to the existing Palivizumab programme, respectively. The scope of the intervention programme however will depend on the purchasing price at when these pharmaceuticals are made available.

## Data Availability

All non-patient sensitive data and modelling code are available online on David Hodgson's github account (see link)

https://github.com/dchodge/rsv_trans_model

## Acknowledgements

**DH:** Medical Research Council PhD Studentship administered through CoMPLEX University College London.

**JPG:** The National Institute for Health Research (NIHR) through the Collaboration for Leadership in Applied Health Research and Care North Thames at Bart’s Health NHS Trust (NIHR CLAHRC North Thames). This funder had no role in study design, data collection, data analysis, data interpretation, or writing of the report. The views expressed in this article are those of the authors and not necessarily those of the NHS, the NIHR, or the UK Department of Health and Social Care.

**RP:** None

**MB:** The MRC Centre for Global Infectious Disease Analysis (grant MR/R015600/1) and the UK National Institute for Health Research Health Protection Research Unit (NIHR HPRU) in Modelling Methodology (Imperial College London) and Immunisation (London School of Hygiene and Tropical Medicine) in partnership with Public Health England (PHE) (grant HPRU-2012–10080) for funding. The views expressed are those of the authors and not necessarily those of the MRC, the UK National Health Service, the UK National Institute for Health Research, the UK Medical Research Council, the UK Department of Health, or Public Health England.

**KA:** The National Institute for Health Research through the Health Protection Research Unit Immunisation at the London School of Hygiene & Tropical Medicine in partnership with Public Health England. The views expressed are those of the authors and not necessarily those of the UK National Health Service, the UK National Institute for Health Research, the UK Medical Research Council, the UK Department of Health, or Public Health England.

## Author’s contributions

DH, KEA, MB, JPG, RP conceived and designed the study. DH performed the mathematical modelling and cost-effectiveness analysis with interpretations from KEA, MB, JPG, RP. DH, KA, drafted the manuscript with critical revisions from JPG, MB, and RP.

---

**Table 3.**
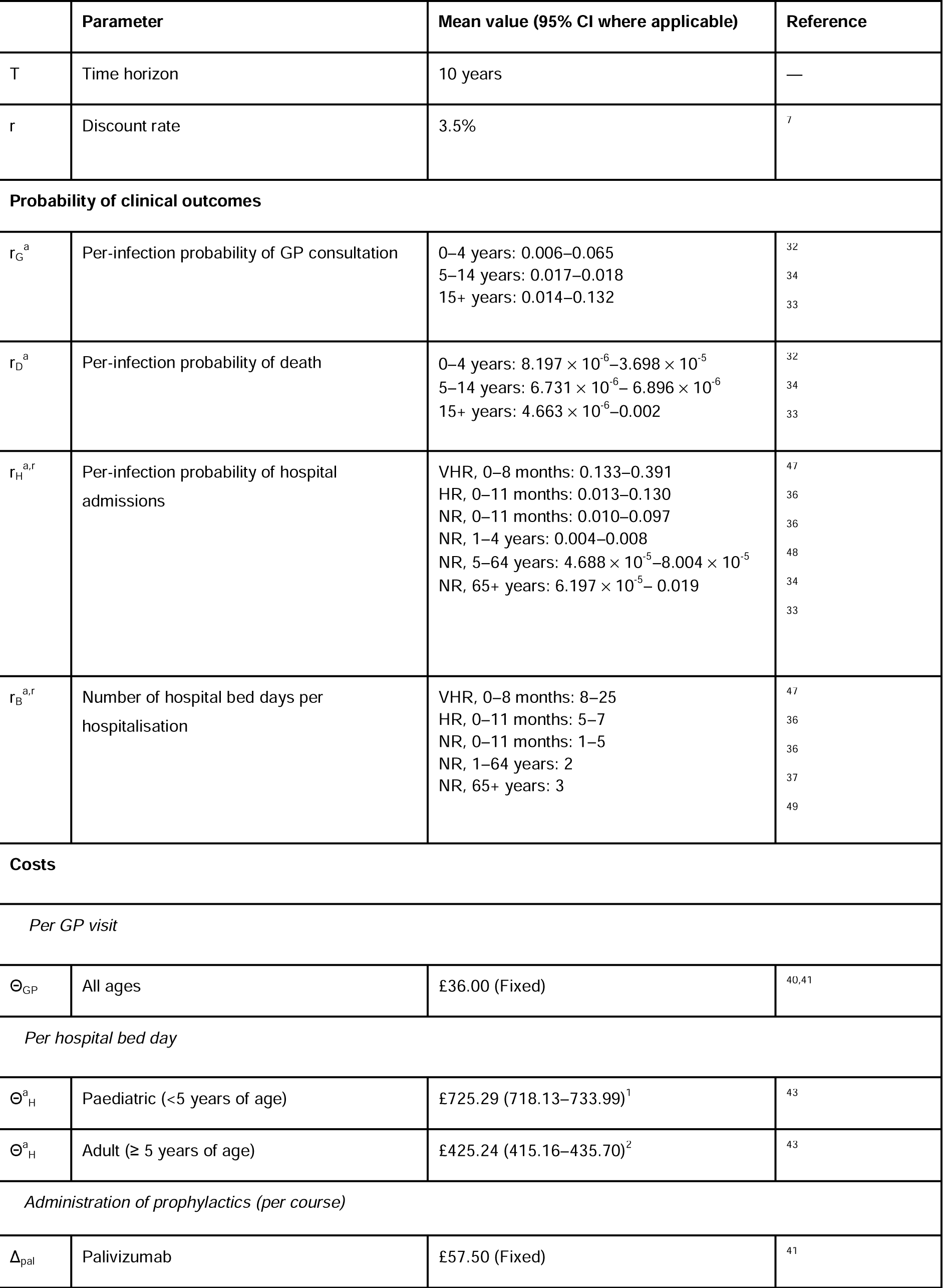
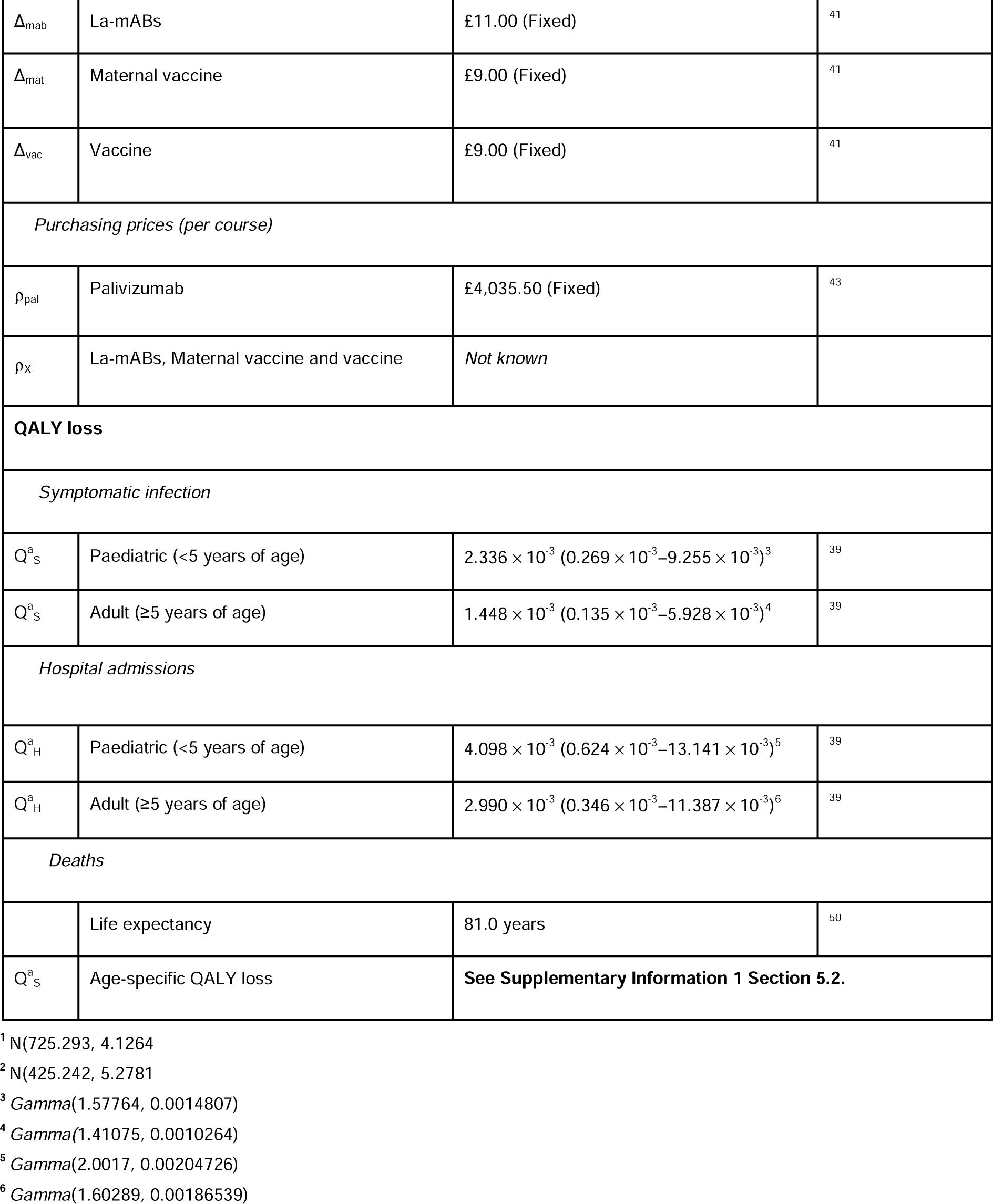
Health and economic parameters used in the cost-effectiveness analysis. Fitted distributions.

|  | Parameter | Mean value (95% CI where applicable) | Reference |
| --- | --- | --- | --- |
| T | Time horizon | 10 years | — |
| r | Discount rate | 3.5% | 7 |
| **Probability of clinical outcomes** |  |  |  |
| $r_G^a$ | Per-infection probability of GP consultation | 0–4 years: 0.006–0.065<br>5–14 years: 0.017–0.018<br>15+ years: 0.014–0.132 | 32<br>34<br>33 |
| $r_D^a$ | Per-infection probability of death | 0–4 years: $8.197 \times 10^{-6}$–$3.698 \times 10^{-5}$<br>5–14 years: $6.731 \times 10^{-6}$–$6.896 \times 10^{-6}$<br>15+ years: $4.663 \times 10^{-6}$–0.002 | 32<br>34<br>33 |
| $r_H^{a,r}$ | Per-infection probability of hospital admissions | VHR, 0–8 months: 0.133–0.391<br>HR, 0–11 months: 0.013–0.130<br>NR, 0–11 months: 0.010–0.097<br>NR, 1–4 years: 0.004–0.008<br>NR, 5–64 years: $4.688 \times 10^{-5}$–$8.004 \times 10^{-5}$<br>NR, 65+ years: $6.197 \times 10^{-5}$–0.019 | 47<br>36<br>36<br>48<br>34<br>33 |
| $r_B^{a,r}$ | Number of hospital bed days per hospitalisation | VHR, 0–8 months: 8–25<br>HR, 0–11 months: 5–7<br>NR, 0–11 months: 1–5<br>NR, 1–64 years: 2<br>NR, 65+ years: 3 | 47<br>36<br>36<br>37<br>49 |
| **Costs** |  |  |  |
| *Per GP visit* |  |  |  |
| $\Theta_{GP}$ | All ages | £36.00 (Fixed) | 40,41 |
| *Per hospital bed day* |  |  |  |
| $\Theta_H^a$ | Paediatric (<5 years of age) | £725.29 (718.13–733.99)<sup>1</sup> | 43 |
| $\Theta_H^a$ | Adult ($\geq 5$ years of age) | £425.24 (415.16–435.70)<sup>2</sup> | 43 |
| *Administration of prophylactics (per course)* |  |  |  |
| $\Delta_{pal}$ | Palivizumab | £57.50 (Fixed) | 41 |
| $\Delta_{\text{mab}}$ | La-mABs | £11.00 (Fixed) | 41 |
| $\Delta_{\text{mat}}$ | Maternal vaccine | £9.00 (Fixed) | 41 |
| $\Delta_{\text{vac}}$ | Vaccine | £9.00 (Fixed) | 41 |
| *Purchasing prices (per course)* |  |  |  |
| $\rho_{\text{pal}}$ | Palivizumab | £4,035.50 (Fixed) | 43 |
| $\rho_{\text{x}}$ | La-mABs, Maternal vaccine and vaccine | *Not known* | |
| **QALY loss** |  |  |  |
| *Symptomatic infection* |  |  |  |
| $Q_{\text{s}}^{\text{a}}$ | Paediatric (<5 years of age) | $2.336 \times 10^{-3} (0.269 \times 10^{-3} - 9.255 \times 10^{-3})^3$ | 39 |
| $Q_{\text{s}}^{\text{a}}$ | Adult ($\geq 5$ years of age) | $1.448 \times 10^{-3} (0.135 \times 10^{-3} - 5.928 \times 10^{-3})^4$ | 39 |
| *Hospital admissions* |  |  |  |
| $Q_{\text{H}}^{\text{a}}$ | Paediatric (<5 years of age) | $4.098 \times 10^{-3} (0.624 \times 10^{-3} - 13.141 \times 10^{-3})^5$ | 39 |
| $Q_{\text{H}}^{\text{a}}$ | Adult ($\geq 5$ years of age) | $2.990 \times 10^{-3} (0.346 \times 10^{-3} - 11.387 \times 10^{-3})^6$ | 39 |
| *Deaths* |  |  |  |
|  | Life expectancy | 81.0 years | 50 |
| $Q_{\text{s}}^{\text{a}}$ | Age-specific QALY loss | **See Supplementary Information 1 Section 5.2.** | |

<sup>1</sup> N(725.293, 4.1264
<sup>2</sup> N(425.242, 5.2781
<sup>3</sup> Gamma(1.57764, 0.0014807)
<sup>4</sup> Gamma(1.41075, 0.0010264)
<sup>5</sup> Gamma(2.0017, 0.00204726)
<sup>6</sup> Gamma(1.60289, 0.00186539)

## References

1 Shi T, McAllister DA, O’Brien KL, et al. Global, regional and national disease burden estimates of acute lower respiratory infections due to respiratory syncytial virus in young children in 2015. Lancet 2017; 390: 946–58.

2 Chapter GB. Respiratory syncytial virus: the green book, chapter 27a. 2015 https://www.gov.uk/government/uploads/system/uploads/attachment_data/file/458469/Green_Book_Chapter_27a_v2_0W.PDF.

3 PATH. RSV vaccine and mAB snapshot. 2019. https://path.azureedge.net/media/documents/RSVsnapshot_2019_04_05_April_High_Resolution.pdf (accessed Aug 20, 2019).

4 Domachowske JB, Khan AA, Esser MT, et al. Safety, Tolerability, and Pharmacokinetics of MEDI8897, an Extended Half-Life Single-Dose Respiratory Syncytial Virus Prefusion F-Targeting Monoclonal Antibody Administered as a Single Dose to Healthy Preterm Infants. Pediatr Infect Dis J 2018; 37: 886–892.

5 Novavax. Prepare™ Trial Topline Results. 2019 https://novavax.com/presentation.show (accessed June 20, 2019).

6 Mazur NI, Higgins D, Nunes MC, et al. The respiratory syncytial virus vaccine landscape: lessons from the graveyard and promising candidates. Lancet Infect Dis 2018; 18: e295–311.

7 NICE. Guide to the methods of technology appraisal. 2013 https://www.nice.org.uk/process/pmg9/resources/guide-to-the-methods-of-technology-appraisal-2013-pdf-2007975843781 (accessed March 14, 2019).

8 Kinyanjui TM, House TA, Kiti MC, Cane PA, Nokes DJ, Medley GF. Vaccine induced herd immunity for control of respiratory syncytial virus disease in a low-income country setting. PLoS One 2015; 10: e0138018.

9 Munywoki PK, Koech DC, Agoti CN, et al. Frequent Asymptomatic Respiratory Syncytial Virus Infections during an Epidemic in a Rural Kenyan Household Cohort. J Infect Dis 2015; 212: 1711–8.

10 Henderson FW, Collier AM, Clyde WA, Denny FW. Respiratory-Sycytial-Virus infections, refinections and immunity: A prospective, longitudinal study in young children. N Engl J Med 1974; 290.

11 Okiro EA, White LJ, Ngama M, Cane PA, Medley GF, Nokes DJ. Duration of shedding of respiratory syncytial virus in a community study of Kenyan children. BMC Infect Dis 2010; 10: 15.

12 Mossong J, Hens N, Jit M, et al. Social contacts and mixing patterns relevant to the spread of infectious diseases. PLoS Med 2008; 5: e74.

13 van Hoek AJ, Andrews N, Campbell H, Amirthalingam G, Edmunds WJ, Miller E. The Social Life of Infants in the Context of Infectious Disease Transmission; Social Contacts and Mixing Patterns of the Very Young. PLoS One 2013; 8: 1–7.

14 Anderson EJ, Carosone-Link P, Yogev R, Yi J, Simões EAF. Effectiveness of Palivizumab in High-risk Infants and Children. 2017. DOI:10.1097/INF.0000000000001533.

15 La BB, Miasojedow Z, and EM, Vihola M. Adaptive Parallel Tempering Algorithm. 2012.

16 Zhao H, Green H, Lackenby A, et al. A new laboratory-based surveillance system (Respiratory Datamart System) for influenza and other respiratory viruses in England: Results and experience from 2009 to 2012. Eurosurveillance 2014; 19: 1–10.

17 Stensballe LG, Ravn H, Kristensen K, Meakins T, Aaby P, Simoes EAF. Seasonal Variation of Maternally Derived Respiratory Syncytial Virus Antibodies and Association with Infant Hospitalizations for Respiratory Syncytial Virus. J Pediatr 2009; 154: 296–9.

18 DeVincenzo JP, Wilkinson T, Vaishnaw A, et al. Viral load drives disease in humans experimentally infected with respiratory syncytial virus. Am J Resp Crit Care 2010; 182: 1305–14.

19 Glezen WP, Paredes A, Allison JE, Taber LH, Frank AL. Risk of respiratory syncytial virus infection for infants from low-income families in relationship to age, sex, ethnic group, and maternal antibody level. J Pediatr 1981; 98: 708–15.

20 Office for National Statistics. Births in England and Wales: 2017. 2018. https://www.ons.gov.uk/peoplepopulationandcommunity/birthsdeathsandmarriages/livebirths/bulletins/birthsummarytablesenglandandwales/2017 (accessed May 14, 2019).

21 Office for National Statistics. Births by parents’ characteristics: 2017. 2019. https://www.ons.gov.uk/peoplepopulationandcommunity/birthsdeathsandmarriages/livebirths/datasets/birthsbyparentscharacteristics (accessed May 14, 2019).

22 Ogilvie MM, Vathenen AS, Radford M, Codd J, Key S. Maternal antibody and respiratroy syncytial virus infection in infancy. J Med Virol 1981; 7: 263–71.

23 Glezen WP, Taber LH, Frank AL, Kasel JA. Risk of primary infection and reinfection with respiratory syncytial virus. Am J Dis Child 1986; 140: 543–6.

24 Ochola R, Sande C, Fegan G, et al. The level and duration of RSV-specific maternal IgG in infants in kilifi Kenya. PLoS One 2009; 4: 4–9.

25 Scott PD, Ochola R, Ngama M, et al. Molecular Analysis of Respiratory Syncytial Virus Reinfections in Infants from Coastal Kenya. J Infect Dis 2006; 193: 59–67.

26 Hall CB, Walsh EE, Long CE, Schnabel KC. Immunity to and frequency of reinfection with respiratory syncytial virus. J Infect Dis 1991; 163: 693–8.

27 ESPID. Tweet from 11th May. 2019. https://twitter.com/jptorrest/status/1127114962839719943 (accessed June 20, 2019).

28 Baguelin M, Hoschler K, Stanford E, Waight P, Hardelid P. Age-Specific Incidence of A/H1N1 2009 Influenza Infection in England from Sequential Antibody Prevalence Data Using Likelihood-Based Estimation. PLoS One 2011; 6: 17074.

29 PHE. Seasonal flu vaccine uptake in GP patients: monthly data, 2018 to 2019 - GOV.UK. 2019. https://www.gov.uk/government/statistics/seasonal-flu-vaccine-uptake-in-gp-patients-monthly-data-2018-to-2019 (accessed June 20, 2019).

30 PHE. Quarterly vaccination coverage statistics for children aged up to five years in the UK (COVER programme)□: October to. 2017 https://assets.publishing.service.gov.uk/government/uploads/system/uploads/attachment_data/file/695475/hpr1118_COVER.pdf (accessed June 20, 2019).

31 Atkins KE, Fitzpatrick MC, Galvani AP, Townsend JP. Cost-Effectiveness of Pertussis Vaccination During Pregnancy in the United States. Am J Epidemiol 2016; 183: 1159–70.

32 Cromer D, van Hoek AJ, Newall AT, Pollard AJ, Jit M. Burden of paediatric respiratory syncytial virus disease and potential effect of different immunisation strategies: a modelling and cost-effectiveness analysis for England. Lancet Public Heal 2017; 2: e367–74.

33 Fleming DM, Taylor RJ, Lustig RL, et al. Modelling estimates of the burden of Respiratory Syncytial virus infection in adults and the elderly in the United Kingdom. BMC Infec Dis 2015; 15: 443.

34 Taylor S, Taylor R, Lustig R, et al. Modelling estimates of the burden of respiratory syncytial virus infection in children in the UK. BMJ Open 2016; 6: e009337.

35 Reeves R, Hardelid P, Gilbert R, Panagiotopoulos N, Minaji M, Pebody R. Using probabilistically linked data to investigate the burden of Respiratory Syncytial Virus (RSV) in children <5 years of age on secondary care in England. Int J Popul Data Sci 2017; 1. DOI:10.23889/ijpds.v1i1.93.

36 Reeves RM, Hardelid P, Panagiotopoulos N, Minaji M, Warburton F, Pebody R. Burden of hospital admissions caused by respiratory syncytial virus (RSV) in infants in England: a data linkage modelling study. J Infect 2019; published online Feb. DOI:10.1016/j.jinf.2019.02.012.

37 Hardelid P, Verfuerden M, McMenamin J, Smyth R, Gilbert R. The contribution of child, family and health service factors to respiratory syncytial virus (RSV) hospital admissions in the first 3 years of life: birth cohort study in Scotland, 2009 to 2015. Eurosurveillance 2019. DOI:10.2807/1560-7917.ES.2019.24.1.1800046.

38 Widmer K, Zhu Y, Williams J V, Griffin MR, Edwards KM, Talbot HK. Rates of hospitalizations for respiratory syncytial virus, human metapneumovirus, and influenza virus in older adults. J Infect Dis 2012; 206: 56–62.

39 Hodgson D, Atkins K, Baguelin M, et al. Estimates for quality of life loss due to RSV (in press). Influ Other Respi Viruses.

40 Hobbs FDR, Bankhead C, Mukhtar T, et al. Clinical workload in UK primary care: a retrospective analysis of 100 million consultations in England, 2007-14. Lancet (London, England) 2016; 387: 2323–30.

41 Curtis LA, Burns A. Unit Costs of Health and Social Care 2018. 2018 DOI:10.22024/UniKent/01.02.70995.

42 Murray J, Bottle A, Sharland M, et al. Risk Factors for Hospital Admission with RSV Bronchiolitis in England: A Population-Based Birth Cohort Study. PLoS One 2014; 9: e89186.

43 NHS Improvement. Reference costs. 2018. https://improvement.nhs.uk/resources/reference-costs/#rc1718 (accessed July 11, 2019).

44 ABHI. NICE, Affordability, and the NHS. 2017 https://www.abhi.org.uk/media/1330/nice-affordability-and-the-nhs.pdf (accessed March 17, 2019).

45 Wolfram Research Inc. Mathematica 11.0.0.0. 2016. http://www.wolfram.com.

46 Reeves RM, Hardelid P, Gilbert R, et al. Epidemiology of laboratory-confirmed respiratory syncytial virus infection in young children in England, 2010-2014: the importance of birth month. Epidemiol Infect 2016; 144: 2049–56.

47 Wang D, Cummins C, Bayliss S, Sandercock J, Burls A. Immunoprophylaxis against respiratory syncytial virus (RSV) with palivizumab in children: A systematic review and economic evaluation. Health Technol Assess (Rockv) 2008; 12. DOI:10.3310/hta12360.

48 Reeves RM, Hardelid P, Gilbert R, Warburton F, Ellis J, Pebody RG. Estimating the burden of respiratory syncytial virus (RSV) on respiratory hospital admissions in children less than five years of age in England, 2007-2012. Influ Other Respi Viruses 2017; 11: 122–9.

49 Widmer K, Zhu Y, Williams J V, Griffin MR, Edwards KM, Talbot HK. Rates of hospitalizations for respiratory syncytial virus, human metapneumovirus, and influenza virus in older adults. J Infect Dis 2012; 206: 56–62.

50 ONS. National life tables, UK - Office for National Statistics. 2018. https://www.ons.gov.uk/peoplepopulationandcommunity/birthsdeathsandmarriages/lifeexpectancies/bulletins/nationallifetablesunitedkingdom/2015to2017 (accessed July 11, 2019).

